# IpsiHand Brain-Computer Interface Therapy Induces Broad Upper Extremity Motor Recovery in Chronic Stroke

**DOI:** 10.1101/2023.08.26.23294320

**Authors:** Nabi Rustamov, Lauren Souders, Lauren Sheehan, Alexandre Carter, Eric C. Leuthardt

**Affiliations:** Division of Neurotechnology, Department of Neurological Surgery, Washington University School of Medicine, St. Louis, MO, United States of America; Center for Innovation in Neuroscience and Technology, Division of Neurotechnology, Washington University School of Medicine, St. Louis, MO, United States of America; Neurolutions, Inc. St. Louis, MO, United States of America; Department of Neurology, Washington University School of Medicine, St. Louis, MO, United States of America; Department of Orthopedic Surgery, Washington University School of Medicine, St. Louis, MO, United States of America; Department of Biomedical Engineering, Washington University in St. Louis, MO, United States of America; Department of Neuroscience, Washington University in St. Louis, MO, United States of America; Department of Mechanical Engineering and Materials Science, Washington University in St. Louis, MO, United States of America

**Author notes:** **Correspondence to: Eric C. Leuthardt MD, FNAI**, Department of Neurological Surgery, Washington University School of Medicine, 660 S. Euclid Avenue, Campus Box 8057, St. Louis, MO 63110, United States of America.

**Keywords:** Brain-Computer Interface, Chronic Stroke, Upper Limb Motor Function Recovery, Theta-Gamma Cross-Frequency Coupling

## Abstract

**Background and Purpose:** Chronic hemiparetic stroke patients have very limited benefits from current therapies. Brain-computer interface (BCI) engaging the unaffected hemisphere has emerged as a promising novel therapeutic approach for chronic stroke rehabilitation. This study investigated the effectiveness of the IpsiHand System, a contralesionally-controlled BCI therapy in chronic stroke patients with impaired upper extremity motor function. We further explored neurophysiological features of motor recovery affected by BCI. We hypothesized that BCI therapy would induce a broad motor recovery in the upper extremity (proximal and distal), and there would be corresponding changes in baseline theta and gamma oscillations, which have been shown to be associated with motor recovery.

**Methods:** Thirty chronic hemiparetic stroke patients performed a therapeutic BCI task for 12 weeks. Motor function assessment data and resting state electroencephalogram (EEG) signals were acquired before initiating BCI therapy and across BCI therapy sessions. The Upper Extremity Fugl-Meyer assessment (UEFM) served as a primary motor outcome assessment tool. Theta-gamma cross-frequency coupling (CFC) was computed and correlated with motor recovery.

**Results:** Chronic stroke patients achieved significant motor improvement with BCI therapy. We found significant improvement in both proximal and distal upper extremity motor function. Importantly, motor function improvement was independent of Botox application. Theta-gamma CFC enhanced bilaterally over the C3 and C4 motor electrodes following BCI therapy. We observed significant positive correlations between motor recovery and theta gamma CFC increase across BCI therapy sessions.

**Conclusions:** BCI therapy resulted in significant motor function improvement across the proximal and distal upper extremities of patients. This therapy was significantly correlated with changes in baseline cortical dynamics, specifically theta-gamma CFC increases in both the right and left motor regions. This may represent rhythm-specific cortical oscillatory mechanism for BCI-driven motor rehabilitation in chronic stroke patients.

## Introduction

Until recently, approaches for chronic stroke rehabilitation provided limited therapeutic benefits. Historically, motor improvements were thought to plateau at three months following stroke, leading to permanent motor deficits ^1–3^. However, recent studies employing Brain-Computer Interface (BCI) strategies for motor rehabilitation have shown that significant functional improvements can be attained even during the chronic phase of stroke ^4–8^. Recently, the United States Food and Drug Administration (FDA) authorized the Neurolutions IpsiHand Upper Extremity Rehabilitation System (IpsiHand System), which is the first BCI therapy for chronic stroke-induced motor impairment. The IpsiHand System uses electroencephalographic (EEG) signals from the unaffected side of the brain to control a distal wearable exoskeleton around the affected hand. Early evidence has shown that regular use of the contralesionally-controlled BCI can facilitate motor recovery in chronic stroke patients ^9–11^.

The contralesional hemisphere has been suggested to play a mechanistic role in post-stroke recovery. Several imaging studies have reported that contralesional activity as well as altered functional connectivity between motor areas in the lesioned and non-lesioned hemispheres are associated with enhanced motor function ^12, 13^. The utilization of the uninjured motor cortex as the signal substrate for BCI motor rehabilitation has further reinforced the positive contribution of the unaffected hemisphere in motor recovery ^10^. A BCI provides very precise temporal coupling of the cortical physiology associated with motor intention from the uninjured hemisphere to the sensory feedback provided to the hand. This BCI-enhanced cortico-sensory coupling is thought to create a Hebbian state to facilitate neuroplastic changes that can improve the brain’s control of the impaired hand. The cortical signals used by the IpsiHand System are low-frequency thalamocortical rhythms in the mu (8-12 Hz) and beta (13-29 Hz) bands that are thought to enable thalamic modulation of the cortex ^14, 15^. Given that these rhythms have a broad cortical distribution ^16^, it remains unclear to what extent the neuroplastic changes induced by the IpsiHand are specific to the hand or whether they have a wider functional effect. Given the emerging nature of this intervention, more data is necessary to define better the magnitude and extent of the benefit provided to these patients.

Here, we aimed to explore the effectiveness of the IpsiHand System for chronic stroke patients with impaired upper extremity motor function. Given that the IpsiHand System is controlled by thalamocortical motor rhythms, we hypothesized that the entire arm would demonstrate a functional improvement. We specifically aimed to assess the global improvement in upper extremity function and also to determine whether the functional benefit was regionally specific (distal limb only) or more general (distal and proximal limb). Further, we assessed whether there was a relationship between the anatomic distribution of functional improvements and the cortical distribution of a previously reported biomarker of theta-gamma coupling that has been shown to track with the functional benefits in chronic stroke rehabilitation ^17^. In aggregate, this study demonstrates the robust benefit that the IpsiHand System provides to the entire upper limb and the associated cortical dynamics.

## Methods

The data are available upon reasonable request to the corresponding author.

### Study Participants

Thirty chronic stroke patients with upper-limb hemiparesis underwent BCI therapy for 12 weeks. Inclusion criteria were stroke at least 6 months prior to this study; unilateral upper extremity weakness. Exclusion criteria were severe visual impairment; severe cognitive impairment quantified by the NIH Stroke Scale; severe aphasia, ataxia, unilateral neglect; joint contractures and impaired tactile or proprioceptive sensation in the affected upper extremity; severe psychiatric disorders; concurrent participation in other stroke studies. Motor function outcomes were primarily assessed with the Upper Extremity Fugl-Meyer assessment (UEFM), which has been validated in a stroke patient population and has high reliability ^18–20^. Secondary motor function outcomes were measured using the Action Research Arm Test (ARAT), Arm Motor Ability Test (AMAT), Motricity Index, Gross Grasp, Modified Ashworth Scale (MAS) at the wrist and elbow. This study was approved by the Institutional Review Board of Washington University School of Medicine in St. Louis. All patients gave written informed consent according to the Declaration of Helsinki.

### BCI System Design

The IpsiHand System consisted of a robotic hand orthosis, EEG amplifier, and wireless EEG headset with six active electrodes (the FDA-approved IpsiHand Upper Extremity Rehabilitation System, Neurolutions, Santa Cruz, CA, USA) (**Fig. 1A**). A touchscreen tablet was connected via bluetooth to the EEG headset to record signals from the brain. Local Wi-Fi network supported communication between the tablet and orthosis. The tablet guided patients through BCI tasks and translated spectral power changes into orthosis control to open and close it in a 3-finger pinch grip. For the BCI task, patients were instructed to open the orthosis with motor imagery of the affected hand or to keep the orthosis closed by resting quietly. The orthosis opened and closed in response to changes in the power of the patient-specific control signal. Subjects who could partially move their affected arm were instructed to allow passive movements by the orthotic device.

**Figure 1.**
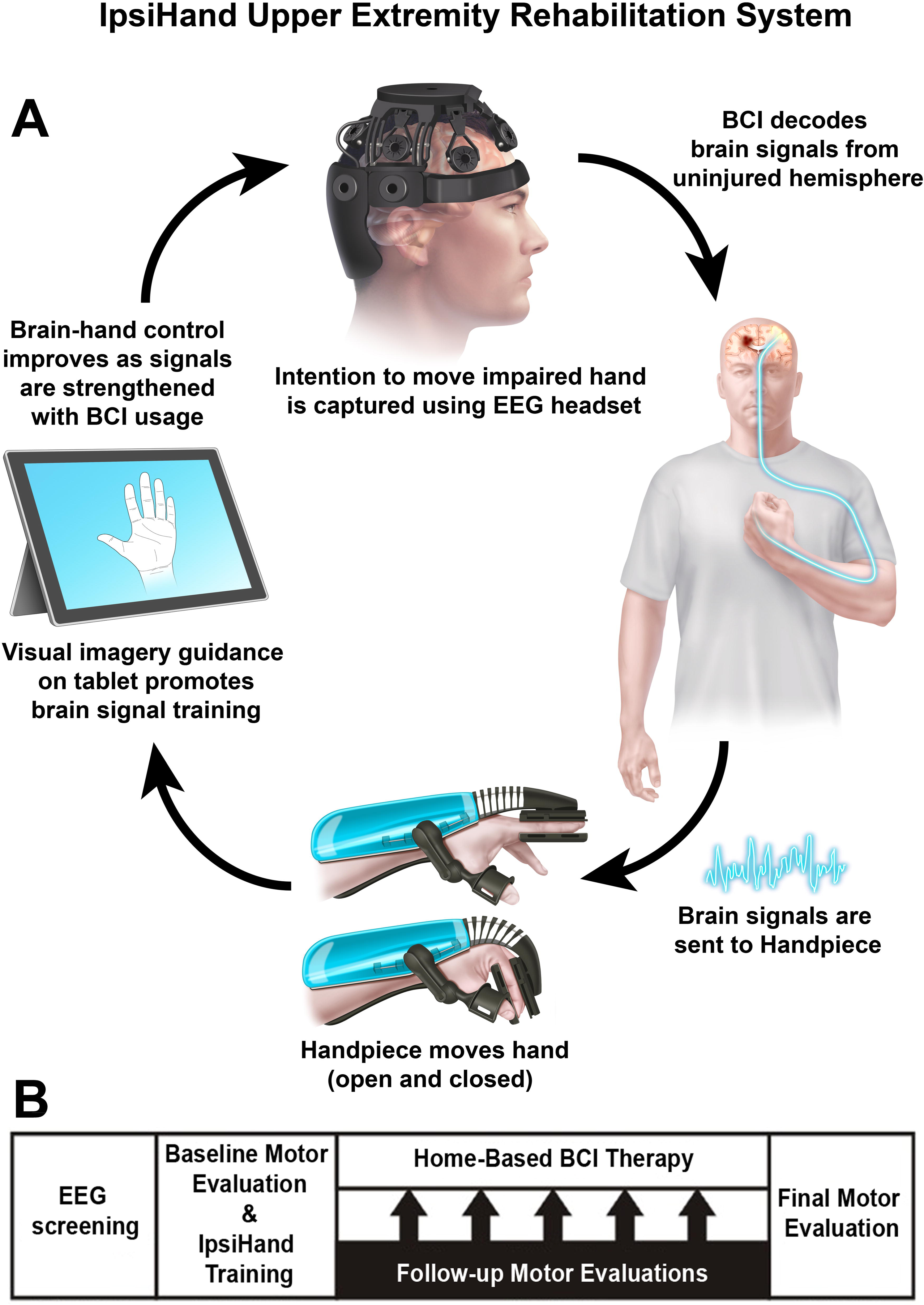
Experimental design. **(A)** BCI System Design. Patients were engaged in motor imagery tasks. Contralesional EEG signals were decoded and translated into commands to open or close the orthosis. Subsequently, the orthosis provided proprioceptive sensory feedback to the patient as they performed motor imagery tasks. **(B)** BCI Therapy Protocol. Patients were screened for their capability to perform the BCI task. Once determined eligible, patients underwent motor assessments prior to commencing the BCI therapy. Daily BCI therapy sessions consisted of one calibration period (extended rest, alternating motor imagery and rest trials) and five BCI therapy runs (motor imagery and rest trials with active orthosis). Fidget periods were incorporated between trials, encouraging patients to blink or make physical adjustments as needed. Motor assessments were conducted every four weeks. After a 12-week therapy, final EEG recordings and motor assessment data were obtained.

### Intervention Protocol

BCI therapy timeline is shown in **Figure 1B**. Patients were first tested for inclusion and exclusion criteria, and the ability to perform the BCI task. During the screening session prior to therapeutic implementation, patients were instructed to perform a series of rest and motor imagery trials. The 1 Hz width frequency band with spectral power modulation best corresponding to the difference between rest and motor trials was selected as the BCI device control signal. The selected control signal was always within the mu (8-12 Hz) or beta (13-29 Hz) canonical frequency band and remained consistent for each patient throughout BCI therapy. Patients with identifiable consistent feature frequency were included in the study. Baseline motor function was evaluated before initiating the therapy by physical and occupational therapists. Research team members then trained patients in the use of the BCI system. Patients were instructed to use the device one hour per day, five days per week, for a total of 12 weeks. Clinicians assessed motor function once every four weeks. The final post-therapy motor assessment was performed after 12 weeks of BCI therapy.

A session of BCI therapy took approximately one hour to complete and consisted of one calibration period and five BCI therapy runs. Pre-therapy calibration was implemented for data quality assurance and for detecting motor imagery activity during the BCI task. During calibration, patients rested quietly then completed a series of task blocks and rest trials. During task blocks, patients were instructed to imagine moving their affected hand. The orthosis did not move during calibration. Following calibration, patients started BCI therapy runs. Each run consisted of 30 motor imagery and 30 rest trials. Trial order was randomized, and three seconds of “fidget” periods were included between each eight-second trial. The “fidget” periods encouraged patients to blink or make physical adjustments. After the completion of BCI therapy run, the system paused to allow patients to rest before continuing with their therapy. Resting state EEG data from pre-task calibration sessions were saved to a remote server for further analysis.

### Electroencephalogram recording and processing

EEG was recorded by means of six wireless dry electrodes (F3, F4, C3, C4, P3 and P4) mounted on the EEG headset in an International 10–20 System (Neurolutions, Santa Cruz, CA, USA). EEG was sampled at 300 Hz with a ground electrode placed on the forehead. Electrode impedance was kept below 10 kΩ. EEG data collected during the pre-therapy calibration rest period were prepared for analysis across BCI therapy sessions (Pre-BCI, Post-BCI 4^th^, 8^th^ and 12^th^ week conditions). The raw EEG data were preprocessed in MATLAB environment (Mathworks, Natick, MA, USA). Resting state EEG data for each condition were five minutes long. EEG recordings were band-pass filtered between 1 and 100 Hz using a finite impulse response (FIR) filter. The 60 Hz notch filter was applied to remove environmental noise. Infomax independent component analysis (ICA) was applied for artifact attenuation ^21^. Independent components found to reflect eye blinks, lateral eye movements, muscle-related and cardiac artifacts were removed. EEG data were common average re-referenced.

### Cross-Frequency Coupling

To calculate theta-gamma CFC, first, raw signal was band-pass filtered in the theta and gamma frequency bands. A Hilbert transform was then applied to obtain the complex-valued analytic signal. Estimates of theta band phase and gamma band amplitude were extracted from the analytic signal. The coupling between theta band (4-8 Hz) phase and gamma band (65-100 Hz) amplitude was quantified using the Mean Vector Length (MVL) approach, originally described in Canolty et al., 2006 ^22^. MVL approach allowed to estimate whether the power at gamma frequencies fluctuated systematically with the phase of the theta frequency, i.e., CFC. Theta-gamma CFC was computed across BCI therapy sessions and compared between conditions.

### Statistical analyses

Changes in motor assessment scores with BCI therapy were assessed in a repeated-measures Analysis of Variance (ANOVA) with within-subjects factor *Session* (Pre-BCI, Post-BCI 4^th^, 8^th^ and 12^th^ week). Planned contrasts were then used to test *a priori hypotheses* and decompose significant effects of BCI therapy. A nonparametric permutation test was performed to examine the differences in mean CFC values following BCI therapy. The electrode labels for each patient were randomly permuted between conditions and the resulting data were used to compute a permutation t-statistic spatiotemporal sensor map for CFC ^23–25^. Correlation analyses were performed between motor assessment scores and theta-gamma CFC values. We used a nonparametric Spearman rank correlation to avoid imposing a model assuming linear relation between variables ^26, 27^. Correlation coefficients were calculated by comparing motor score and CFC value changes across BCI therapy sessions. All statistical tests were two-tailed with a significance level of 0.05 and the P-values were adjusted using a Bonferroni correction.

## Results

### Patient characteristics

Patient disposition and characteristics are shown in **Suppl. Figure 1** and **Suppl. Table 1**, respectively. The age was 58.7 ± 11.5 years (mean ± standard deviation); 9 females, 21 males.

### Motor Assessment Scores

The UEFM served as a primary motor outcome assessment tool. All patients achieved an increase in UEFM score following BCI therapy. The mean increase in UEFM score was 7.77 points which exceeded the minimal clinically significant difference (MCID) threshold of 5.25 points increase ^28^. A total of 21 out of the 30 patients reached the MCID.

Primary and secondary motor assessment scores across BCI therapy sessions were presented in **Figures 2 and 3**. **UEFM:** the main effect of session was significant, F(3,87) = 32.06, p = 0.001. Planned contrasts revealed significant increases in UEFM scores. **ARAT:** the main effect of session was significant, F(3,87) = 15.91, p = 0.001. Planned contrasts revealed significant increases in ARAT scores. **AMAT:** the main effect of session was significant, F(3,87) = 26.43, p = 0.001. Planned contrasts revealed significant increases in AMAT scores. **Motricity Index:** the main effect of session was significant, F(3,87) = 18.66, p = 0.001. Planned contrasts revealed significant increases in motricity index scores. **Gross Grasp:** the main effect of session was significant, F(3,87) = 6.14, p = 0.001. Planned contrasts revealed significant increases in motricity index scores. **Elbow and Wrist MAS:** the main effect of session did not prove significant, F(3,87) = 2.28 and 0.42, p = 0.09 and 0.74, respectively, excluding significant changes across BCI therapy sessions.

**Figure 2.**
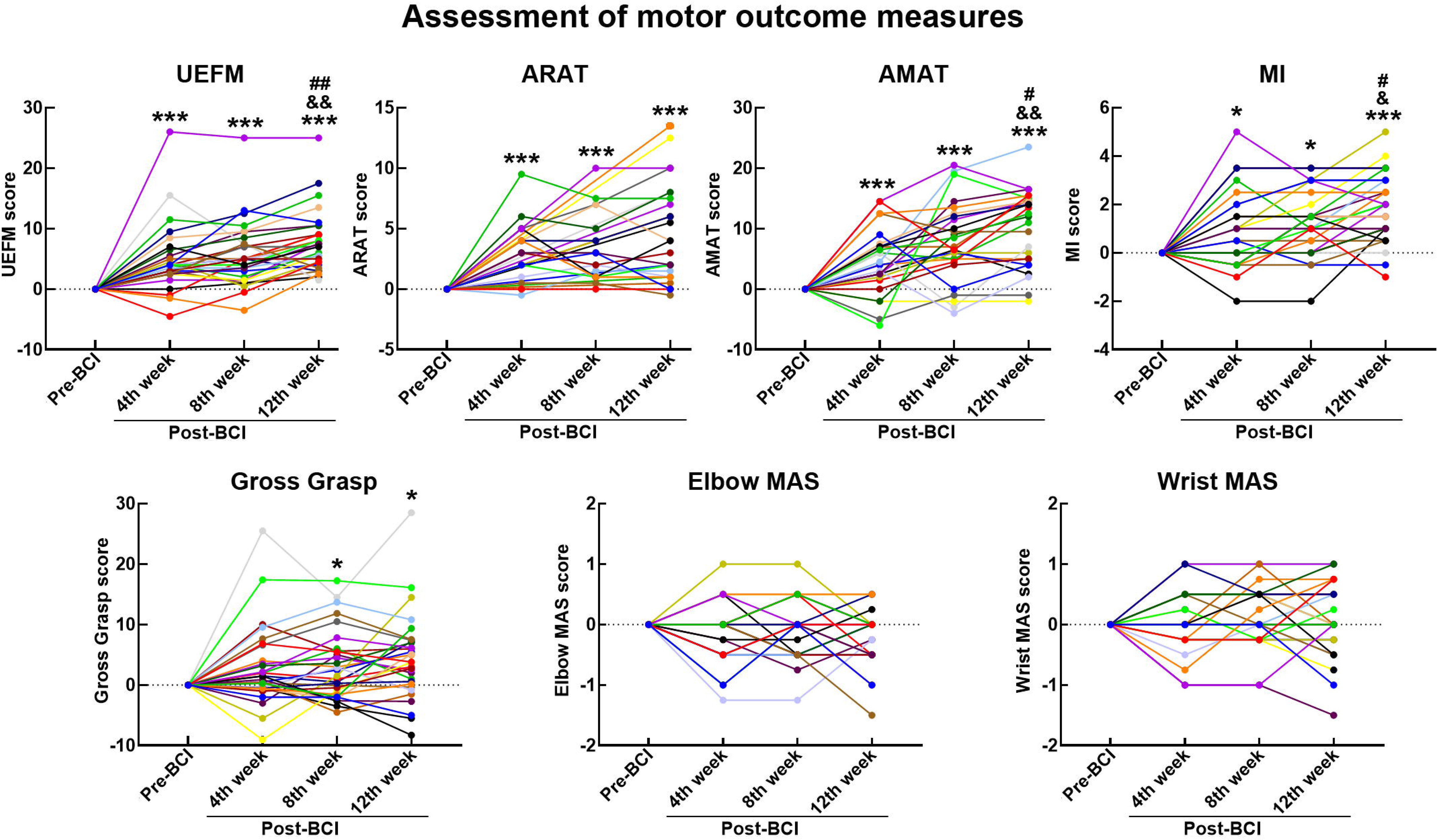
Motor assessment scores. Motor scores across BCI therapy sessions. Y-axis, motor score; X-axis, BCI therapy sessions. UEFM: Upper Extremity Fugl-Meyer; ARAT: Action Research Arm Test; AMAT: Arm Motor Ability Test; MAS: Modified Ashworth Scale. Patients are depicted in different colors. Significance levels were based on the Pairwise Comparisons in ANOVA (Bonferroni corrected). *, **, *** symbols: pC≤C0.05, 0.01 and 0.001 for Pre-BCI vs. Post-BCI 4^th^, 8^th^ and 12^th^ week contrasts; &, && symbols: pC≤C0.05 and 0.01 for Post-BCI 4^th^ vs. 12^th^ week contrasts; #, ## symbols: pC≤C0.05 and 0.01 for and Post-BCI 8^th^ vs. 12^th^ week contrast.

**Figure 3.**
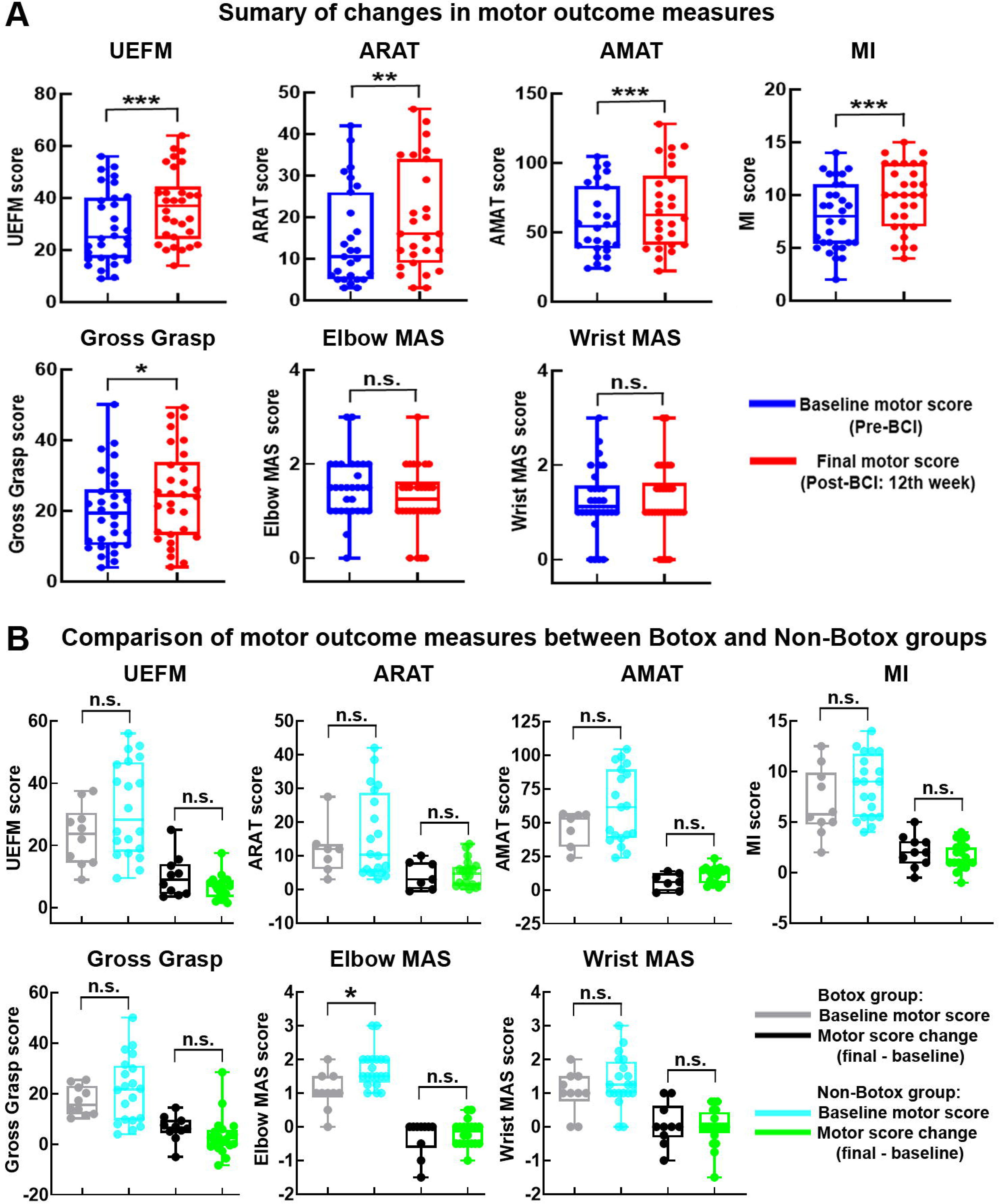
Summary of motor recovery measures. **(A)** Baseline and final motor scores. **(B)** Botox and Non-Botox groups. Baseline motor scores and final motor score changes. Y-axis, motor score; X-axis, BCI therapy sessions. UEFM: Upper Extremity Fugl-Meyer; ARAT: Action Research Arm Test; AMAT: Arm Motor Ability Test; MAS: Modified Ashworth Scale. *, **, *** symbols: pC≤C0.05, 0.01 and 0.001; n.s.: not statistically significant.

### Upper Extremity Fugl-Meyer Section Scores

We evaluated changes in UEFM section scores following BCI therapy (see **Suppl. Table 2** for UEFM section description). The results are summarized in **Figure 4**. **Section A 1** (triceps/ biceps reflex): the main effect of session was not significant, F(3,87) = 2.43, p = 0.07, excluding significant changes across BCI therapy sessions. **Sections A2, A3 and A4** (proximal upper extremity): the main effect of session was significant, F(3,87) = 16.70, 3.48 and 6.84, p = 0.001, 0.02 and 0.001, respectively. Planned contrasts revealed significant increases in UEFM section scores. **Sections B and C** (distal upper extremity): the main effect of session was significant, F(3,87) = 9.35 and 12.16, p = 0.001 and 0.001, respectively. Planned contrasts revealed significant increases in UEFM section scores. **Section D** (whole limb coordination): the main effect of session was significant, F(3,87) = 6.60, p < 0.001. Planned contrasts revealed significant increases in UEFM section scores.

**Figure 4.**
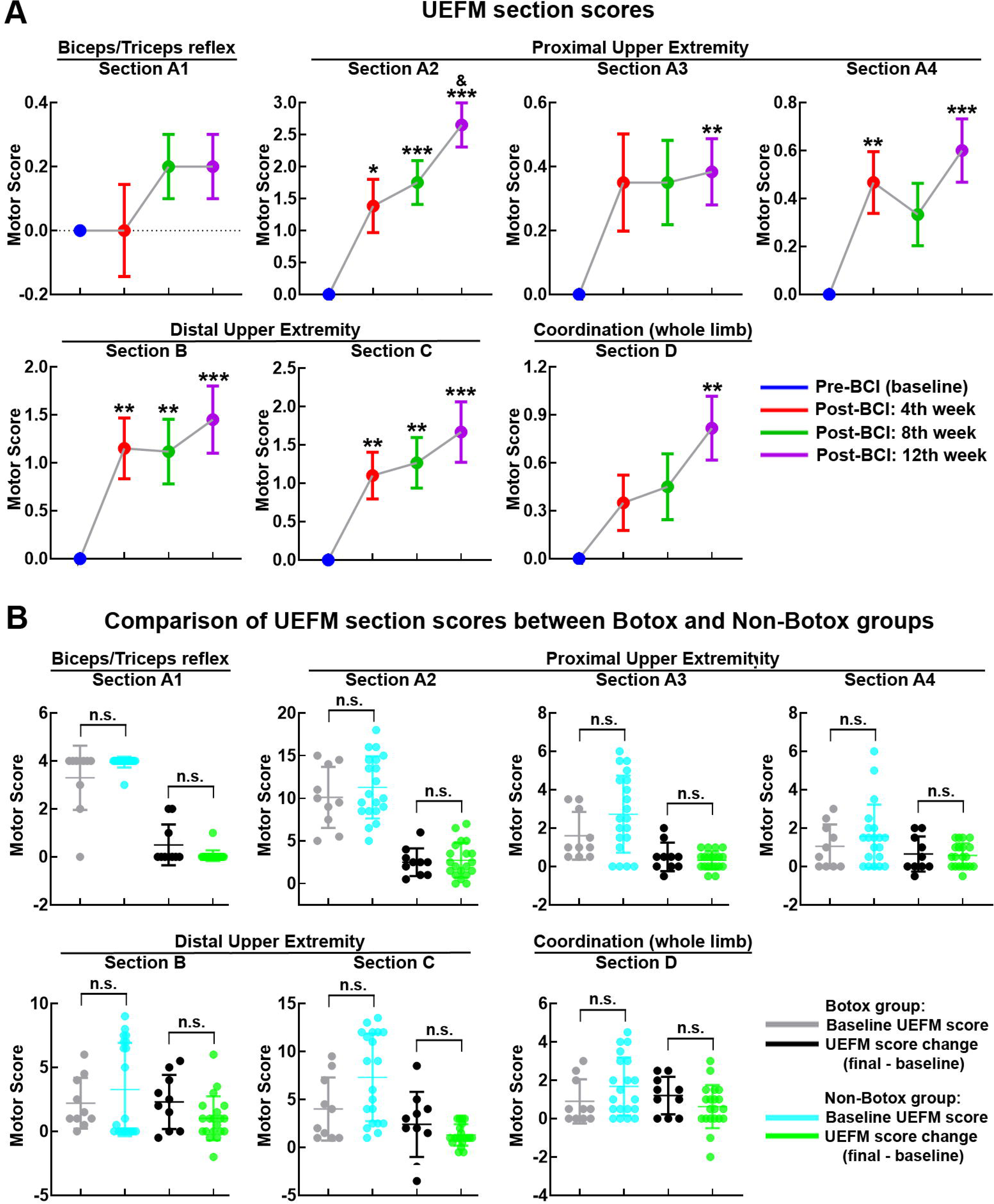
Section scores of UEFM. **(A)** UEFM section scores across BCI therapy sessions. **(B)** Botox and Non-Botox groups. Baseline UEFM section scores and final UEFM section score changes. Y-axis, motor score; X-axis, BCI therapy sessions. UEFM: Upper Extremity Fugl-Meyer. *, **, *** symbols: pC≤C0.05, 0.01 and 0.001 for Pre-BCI vs. Post-BCI 4^th^, 8^th^ and 12^th^ week contrasts; & symbol: pC≤C0.05 for Post-BCI 4^th^ vs. 12^th^ week contrast; n.s.: not statistically significant.

### Comparison of motor recovery between Botox and Non-Botox groups

Both Botox and Non-Botox patients achieved greater than MCID gains in UEFM scores (10.05 and 6.63 points gain, respectively). The proportion of patients in each group who met the MCID for UEFM was similar: 70% (7/10) of Botox patients and 65% (13/20) of Non-Botox patients. Further statistical analyses were performed to assess if the influence of Botox application on motor recovery following BCI therapy was significant. Pre-BCI baseline motor scores and final motor score change (motor score Post-BCI 12^th^ week - Pre-BCI baseline motor score) were compared between Botox (n = 10) and Non-Botox groups (n = 20) (**Figures 3B and 4B**). The differences in baseline motor scores and final motor score change between groups were not statistically significant (all p’s > 0.05), except baseline Elbow MAS score. However, although baseline Elbow MAS score was significantly lower in Botox group compared with Non-Botox group (p = 0.03), groups did not significantly differ in final Elbow MAS score change (p = 0.42) (**Figure 3B**). The Botox and Non-Botox groups were also compared for the UEFM section scores. The results presented in **Figure 4B** indicate that neither baseline motor scores nor final motor score changes were statistically different between groups for any section of the UEFM (all p’s > 0.05).

### Modulation of brain activity

#### Theta-gamma cross-frequency coupling

**Figure 5A** depicts topographic representation of mean theta-gamma CFC values across BCI therapy sessions (Pre-BCI, Post-BCI 4^th^, 8^th^ and 12 week). We compared theta-gamma CFC values between Pre-BCI baseline and Post-BCI sessions as measured over all the electrode sites (**Figure 5B**). During the earlier post-treatment session (Post-BCI 4^th^ week), theta-gamma CFC increased at the C3 and C4 motor electrodes. This was in the expected direction but did not reach significance (both p’s > 0.05). The subsequent Post-BCI sessions were associated with significant bilateral enhancement of theta-gamma CFC over the motor electrodes (Post-BCI 8^th^ week: significant effects at the C3 and C4 electrodes, both p’s < 0.05; Post-BCI 12^th^ week: significant effects at the C3 and C4 electrodes, both p’s < 0.05). Post-BCI 12^th^ week vs. Post-BCI 8^th^ week contrast did not reveal significant difference in theta-gamma CFC (all p’s > 0.05). Theta-gamma CFC over the frontal and parietal electrodes was not significantly modulated throughout BCI therapy (all p’s > 0.05).

**Figure 5.**
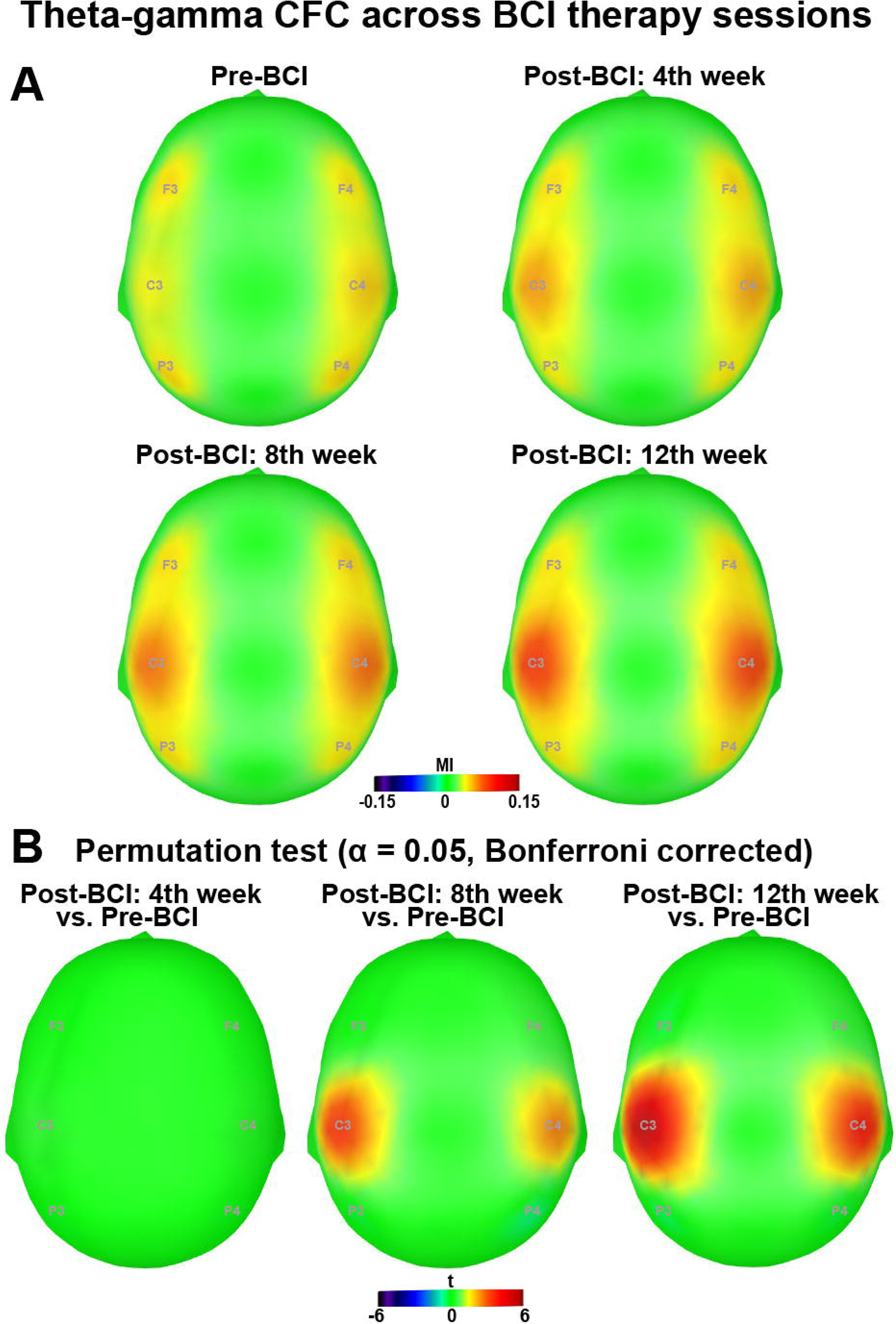
Topographic representation of mean theta-gamma CFC values. **(A)** Theta-gamma CFC values across BCI therapy sessions. **(B)** Electrode level t maps of the comparison between conditions as assessed by nonparametric permutation tests. Only electrodes whose t statistic exceeded a critical threshold of P ≤ 0.05 (two-tailed, Bonferroni corrected) were retained. For the electrodes not showing significant effects, t values were set to zero. Higher values are depicted by red color. CFC: Cross-Frequency Coupling; BCI: Brain-Computer Interface; MI: Modulation Index.

### Correlations between motor recovery and theta-gamma cross-frequency coupling

Correlations between motor recovery and theta-gamma CFC change at the C3 and C4 electrodes across BCI therapy sessions relative to baseline are shown in **Figure 6**. UEFM, AMAT and Motricity Index score changes showed significant positive correlations with theta-gamma CFC change. ARAT, Gross Grasp, Elbow and Wrist MAS score changes did not correlate significantly with theta-gamma CFC change. We also conducted correlation analyses for UEFM section scores (**Suppl. Fig. 2**). The gains in both proximal and distal UEFM section scores correlated significantly with theta-gamma CFC change at the C3 and C4 electrodes.

**Figure 6.**
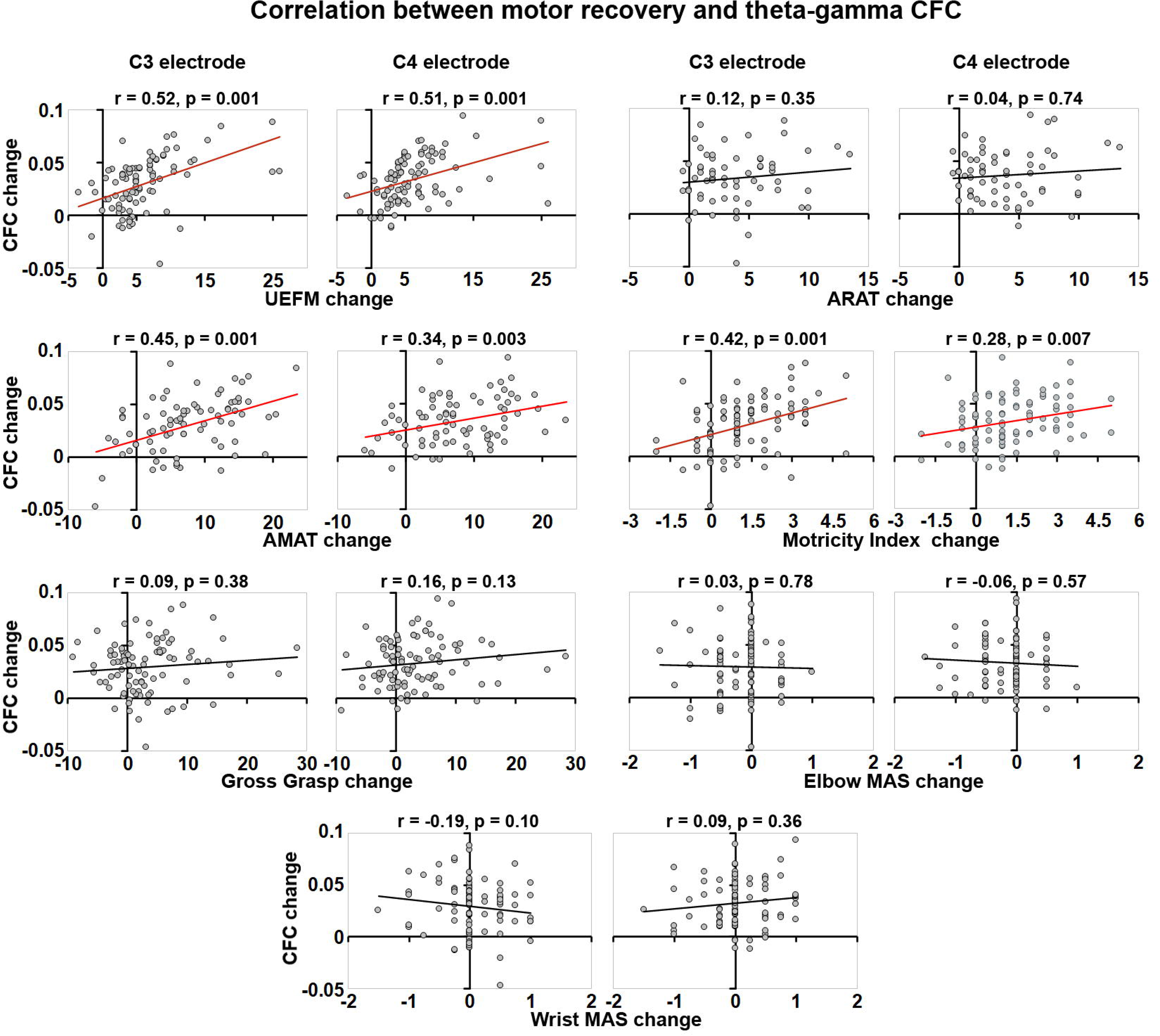
Correlation between motor recovery and theta-gamma CFC modulation. Spearman rank correlations were run between changes in motor scores and theta-gamma CFC values across BCI therapy sessions relative to Pre-BCI baseline. Significant and non-significant correlations were depicted by red and black color, respectively. Y-axis, CFC change; X-axis, motor score change. CFC: Cross-Frequency Coupling; UEFM: Upper Extremity Fugl-Meyer; ARAT: Action Research Arm Test; AMAT: Arm Motor Ability Test; MAS: modified Ashworth Scale.

## Discussion

This study provides compelling evidence demonstrating that the use of a contralesionally-controlled BCI for chronic stroke therapy, the IpsiHand System, induced clinically significant motor function improvement. Despite being more than six months post-stroke, patients with chronic stroke achieved a significant increase in UEFM score. Importantly, we found improvement in both proximal and distal upper extremity section scores of UEFM. Additionally, improvements were observed in secondary measures of motor function. A significant increase was evident in ARAT, AMAT, Motricity Index and Gross Grasp scores, which collectively assess proximal and distal upper extremity motor function. Conversely, the measurements of muscle spasticity, such as Elbow and Wrist MAS, did not exhibit significant changes following BCI intervention. These findings further corroborate previous evidence, reinforcing the notion that BCI-controlled rehabilitation systems can facilitate motor recovery ^5, 6, 8^. However, this study was unique in utilizing the unaffected hemisphere in a BCI rehabilitation system and demonstrating the benefit for improvement is widespread in the affected limb, setting it apart from previous research. Our findings provide support for the engagement of the unaffected hemisphere as an important substrate for motor rehabilitation.

In this study, both Botox and Non-Botox groups achieved greater than MCID gains in UEFM scores. In addition, the proportion of patients in each group who met the MCID for UEFM was nearly the same. BCI therapy provided improvement in both proximal and distal upper extremity section scores of UEFM. The gains for each specific UEFM section were also not significantly different between Botox and Non-Botox groups. These results indicate that Botox application did not have a statistically significant impact on the benefit provided by the BCI therapy. The use of Botox to reduce spasticity may have functional effects that both facilitate and impair motor recovery. In a previous study, Botox application for post-stroke upper limb spasticity did not improve upper limb motor function, including activities such as grasping and reaching ^29^. Botox has also been associated with increased motor impairment, including weakness of injected muscles ^30^. Thus, while reducing muscle tone can enhance finer movements, it is important to acknowledge that it may also decrease strength and hinder overall motor function. Regardless, the presence of Botox did not facilitate or hinder the effect of the IpsiHand System’s functional benefits.

In addition to the functional gains associated with the use of the IpsiHand System, we also evaluated whether there were concomitant cortical electrophysiologic changes associated with these motor improvements. Given prior work that showed theta-gamma CFC was associated with BCI rehabilitation ^17^, we further evaluated the dynamic changes in theta-gamma CFC throughout BCI therapy to evaluate the neurophysiological mechanisms underlying motor recovery. In previous studies, the interaction between neuronal oscillations at different frequency bands, i.e., CFC, has been observed in various brain regions, including the hippocampus ^31, 32^, subcortical nuclei ^33, 34^ and neocortex ^22, 35^. While the exact functional significance of CFC remains uncertain, it is believed to play a crucial role in coordinating neuronal activity across brain regions ^36, 37^. A previous study has demonstrated the association between enhanced theta-gamma coupling over motor regions and motor recovery in chronic stroke patients. This effect was driven by amplified synchronization of underlying resting theta and gamma oscillations rather than changes in their power magnitude ^17^. During the learning of item-context associations, theta-gamma coupling showed positive correlation with behavioral performance accuracy suggesting its involvement in learning process ^38^. Theta-gamma coupling has been shown to emerge in the primary motor cortex when gamma-aminobutyric acidergic (GABAergic) activity is inhibited ^39^. Reduced GABAergic activity in the primary motor cortex represents a central mechanism for motor plasticity ^40, 41^. Taken together, it can be inferred that enhanced synchronization of theta and gamma oscillations serves as an important hallmark of motor learning. A key electrophysiologic finding of our study was the bilateral enhancement of coupling between theta and gamma oscillations over the motor areas following BCI therapy. This effect showed a significant positive correlation with both proximal and distal upper limb motor improvement. Additionally, our study revealed that theta-gamma CFC progressively increased in later therapy sessions compared to the earlier ones. These findings suggest that theta-gamma coupling may play a mechanistic role in motor recovery. This aligns with previous research indicating the involvement of theta-gamma interaction in spatial and motor learning processes ^42–44^. Our findings also provide support for previous studies highlighting the involvement of the primary sensory-motor cortex in learning-related processes ^43, 45–47^.

This study was conducted with the assumption that motor deficits would remain stable in the chronic stage of stroke. Thus, we did not have a BCI control group. Indeed, previous research has shown limited improvement in motor deficits during the chronic stage of stroke ^2, 3, 48, 49^. Furthermore, in a separate study investigating motor rehabilitation in stroke patients, sham BCI therapy failed to facilitate motor recovery ^6^. Consequently, we primarily attribute the observed improvement in motor function and associated electrophysiological findings in this study to the BCI intervention. It is important to note that we cannot currently determine whether enhanced theta-gamma coupling is specific to BCI techniques or if it represents a broader phenomenon associated with other methods utilized for chronic stroke rehabilitation.

In conclusion, in this study we aimed to investigate the effectiveness of a contralesionally-controlled BCI therapy (IpsiHand System) in chronic stroke patients with impaired upper extremity motor function. Patients achieved significant improvement in both proximal and distal upper extremity motor function, which was independent of Botox application. Motor recovery was paralleled with enhanced theta and gamma coupling over the motor regions, which may represent rhythm-specific cortical oscillatory mechanisms for BCI-driven motor rehabilitation. This study represents a crucial advancement in the development and translation of BCI therapy protocols designed for chronic stroke rehabilitation. Our findings warrant large-scale randomized controlled trials to further establish the effectiveness of BCI-driven motor rehabilitation in chronic stroke.

## Data Availability

The data are available upon reasonable request to the corresponding author.

## Acknowledgements

The authors thank study participants for their time and effort.

## Funding

This work was supported by National Institutes of Health (NIH) R21NS102696 (E.C.L. and A.C.) and National Institute of Biomedical Imaging and Bioengineering (NIBIB) P41-EB018783 (E.C.L.).

## Disclosures

E.C.L. owns stock in Neurolutions, Inner Cosmos and Sora Neuroscience. Washington University owns stock in Neurolutions. This work and E.C.L. have had their conflict of interest rigorously evaluated and managed throughout this study and with the creation of this manuscript. Other authors have nothing to disclose.

## Notes

### Competing Interest Statement

The authors have declared no competing interest.

### Clinical Trial

NCT03611855

### Author Declarations

Ethics committee/IRB of Washington University School of Medicine in St. Louis gave ethical approval for this work.

