## Supplemental Figures and Tables for "IpsiHand Brain-Computer Interface Therapy Induces Broad Upper Extremity Motor Recovery in Chronic Stroke"

**Supplemental Material:**

Supplementary Figure 1. Patient recruitment flowchart.

Supplementary Figure 2. Correlation between UEFM section scores and theta-gamma CFC.

Supplementary Table 1: Patient characteristics.

Supplementary Table 2: Upper Extremity Fugl-Meyer Section Description.

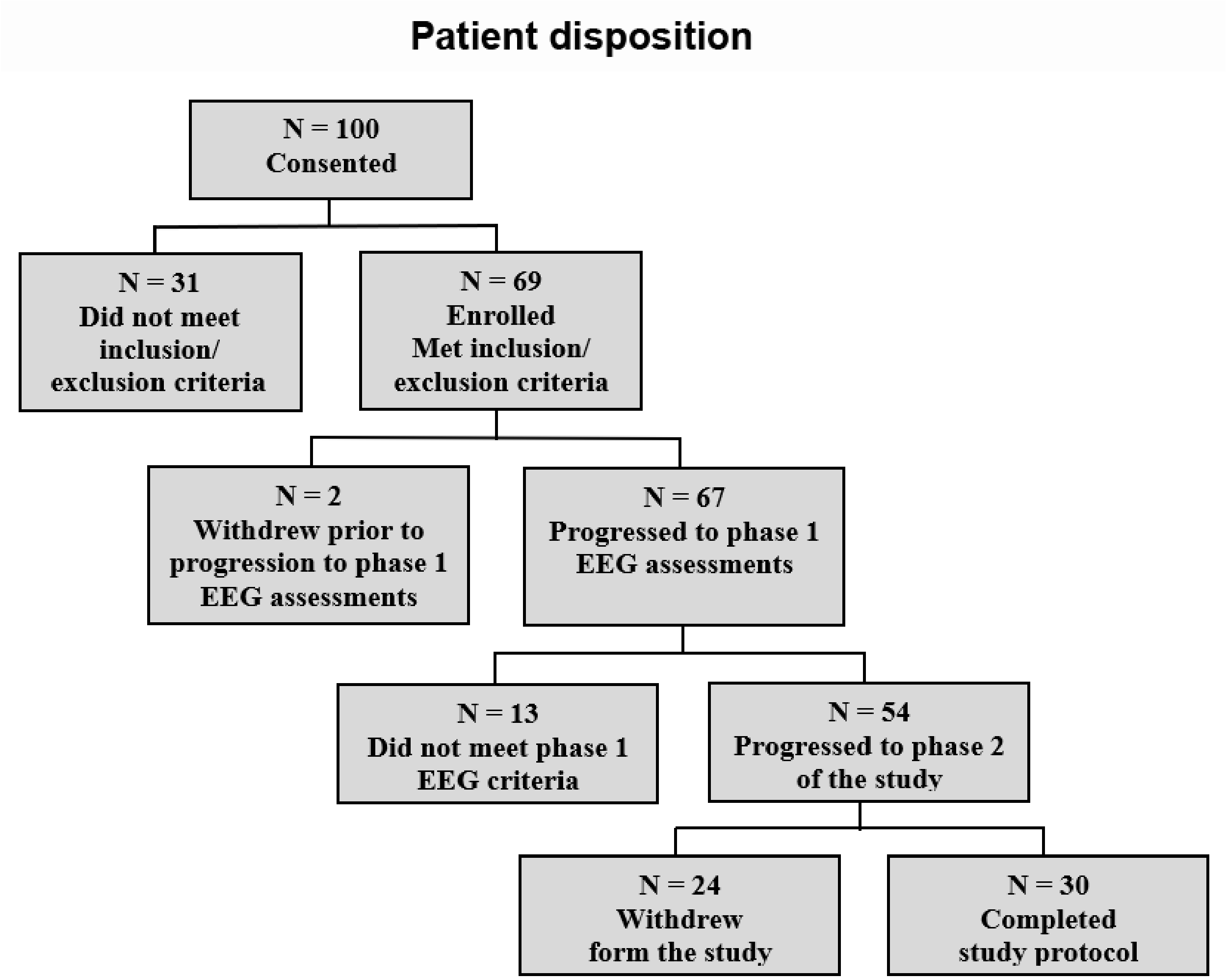


**Supplementary Figure 1. Patient recruitment flowchart.** One hundred patients with chronic stroke consented to participate in the study. Thirty-one patients did not meet the eligibility criteria. Sixty-nine patients were enrolled in the study. Thirty patients completed the study.

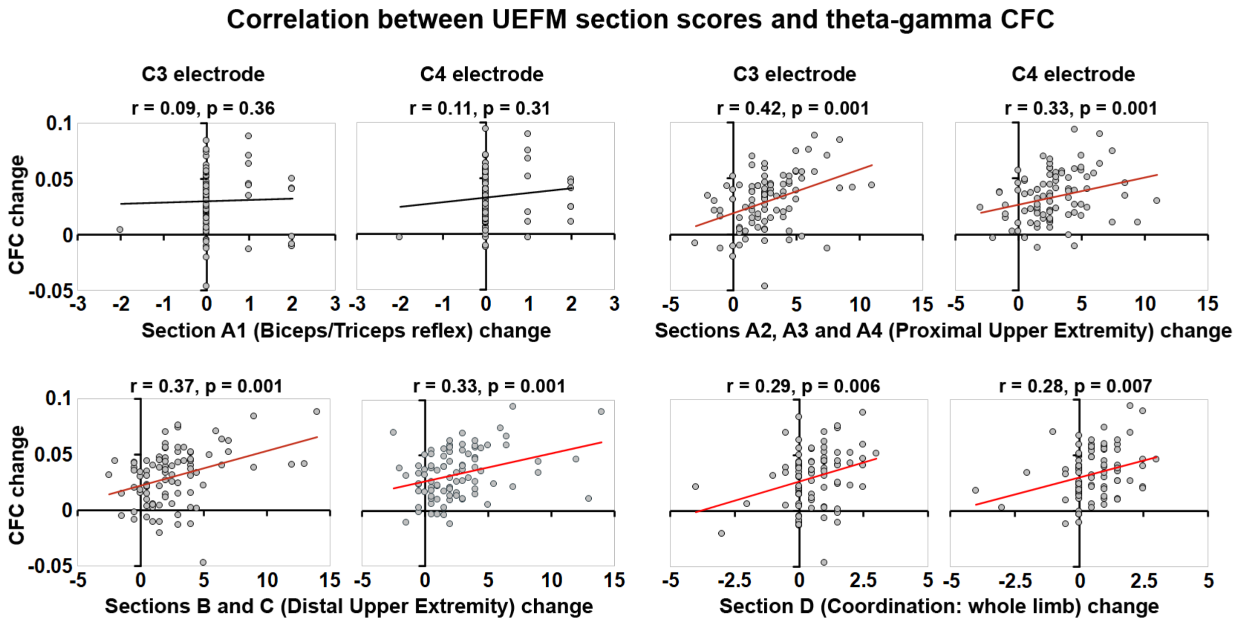


**Supplementary Figure 2. Correlation between UEFM section scores and theta-gamma CFC.** Spearman rank correlations were run between changes in UEFM section scores and theta-gamma CFC values across BCI therapy sessions relative to Pre-BCI baseline. Significant and non-significant correlations were depicted by red and black color, respectively. Y-axis, CFC change; X-axis, UEFM section scores change. UEFM: Upper Extremity Fugl-Meyer; CFC: Cross-Frequency Coupling.

**Supplementary Table 1: Patient characteristics.**

| Patient | Diagnosis (location) | Affected UE / Hand Dominance | Time Post-Stroke (months) | Botox Usage (Yes / No) |
|---|---|---|---|---|
| 1 | Right Hemispheric CVA | LUE/ Right | 44 | Yes |
| 2 | Right Hemispheric CVA | LUE/ Right | 153 | No |
| 3 | Left Hemispheric CVA | RUE/ Right | 20 | Yes |
| 4 | Left Hemispheric CVA | RUE/ Right | 9 | Yes |
| 5 | Left Hemispheric CVA | RUE/ Right | 19 | No |
| 6 | Basal ganglia CVA | LUE/ Right | 61 | No |
| 7 | Left Hemispheric CVA | RUE/ Right | 372 | Yes |
| 8 | Left Hemispheric CVA | RUE/ Right | 24 | No |
| 9 | Left Hemispheric CVA | RUE/ Right | 20 | No |
| 10 | Right Hemispheric CVA | LUE/ Right | 22 | No |
| 11 | Left Hemispheric CVA | RUE/ Right | 20 | Yes |
| 12 | Left Hemispheric CVA | RUE/ Right | 34 | Yes |
| 13 | Right Hemispheric CVA | LUE/ Right | 253 | No |
| 14 | Right Hemispheric CVA | LUE/ Right | 38 | Yes |
| 15 | Left Hemispheric CVA | RUE/ Right | 65 | No |
| 16 | Right Hemispheric CVA | LUE/ Right | 34 | No |
| 17 | Right Hemispheric CVA | LUE/ Right | 122 | No |
| 18 | Left Hemispheric CVA | RUE/ Right | 156 | No |
| 19 | Left Hemispheric CVA | RUE/ Right | 30 | No |
| 20 | Right Hemispheric CVA | LUE/ Right | 96 | Yes |
| 21 | Right Hemispheric CVA | LUE/ Left | 35 | Yes |
| 22 | Left Hemispheric CVA | RUE/ Right | 29 | No |
| 23 | Left Hemispheric CVA | RUE/ Right | 16 | No |
| 24 | Left Basilar CVA | RUE/ Right | 28 | Yes |
| 25 | Right Hemispheric CVA | LUE/ Right | 12 | No |
| 26 | Left Hemispheric CVA | RUE/ Right | 103 | No |
| 27 | Right Hemispheric CVA | LUE/ Left | 27 | No |
| 28 | Left Hemispheric CVA | RUE/ Right | 68 | No |
| 29 | Left Hemispheric CVA | RUE/ Right | 108 | No |
| 30 | Right Hemispheric CVA | LUE/ Right | 28 | No |

CVA: cerebrovascular accident; LUE: left upper extremity; RUE: right upper extremity; UE: upper extremity.

### Supplementary Table 2: Upper Extremity Fugl-Meyer Section Description.

| **Upper Extremity Fugl-Meyer Section** | **Description** | **Total Points** |
|---|---|---|
| Section A1: Reflex Activity | Assesses whether a reflex is present in the triceps or biceps of the affected upper extremity. | 4 points |
| Section: A2: Volitional Movement with Synergies (without gravitational help) | Assesses the subject's ability to move volitionally against gravity within typical stroke recovery synergy patterns (hand to opposite knee and up and behind the ear on the affected side). | 18 points |
| Section A3: Volitional Movement Mixing Synergies (without compensation) | Assesses the subject's ability to move in combination patterns (i.e., hand to lumbar spine, shoulder flexion to 90 degrees and pronation and supination) without compensatory movement. | 6 points |
| Section A4: Volitional Movement with Little or No Synergy | Assesses the subject's ability to move in patterns (i.e., shoulder flexion from 90-180 degrees, shoulder abduction and supination/ pronation with elbow at 0 degrees) to determine the ability to move outside of synergistic stroke recovery movement patterns. | 6 points |
| Section A5: Normal Reflex Activity | Only assessed if the subject scores 6 points in Section A4. Assesses a subject's hyperreactivity of the biceps, triceps, and finger flexors. | 2 points |
| Section B: Wrist | Assesses the subject's ability to move the wrist against gravity, against resistance, and assesses the smoothness and fluidity of wrist movement in multiple planes. | 10 points |
| Section C: Hand | Assesses the subject's ability to perform mass flexion and extension as well as multiple grasp patterns against resistance. | 14 points |
| Section D: Coordination/ Speed | Assesses the subject's fluidity and timing for finger to nose movement on the affected side in comparison with the unaffected side. | 6 points |
